# Genetic Susceptibility to Astrovirus Diarrhea in Bangladeshi Infants

**DOI:** 10.1101/2023.07.03.23292186

**Authors:** Laura Chen, Rebecca M. Munday, Rashidul Haque, Dylan Duchen, Uma Nayak, Poonum Korpe, Alexander J. Mentzer, Beth D. Kirkpatrick, Genevieve L. Wojcik, William A. Petri, Priya Duggal

**Affiliations:** Department of Epidemiology, Johns Hopkins Bloomberg School of Public Health; Department of Genetic Medicine, Johns Hopkins University School of Medicine; International Center for Diarrhoeal Disease Research, Bangladesh; Center for Public Health Genomics, University of Virginia School of Medicine; Department of Public Health Sciences, University of Virginia School of Medicine; The Wellcome Centre for Human Genetics, University of Oxford; Department of Microbiology and Molecular Genetics, Vaccine Testing Center, Larner College of Medicine, University of Vermont; Department of Medicine, Infectious Diseases and International Health, University of Virginia School of Medicine

## Abstract

Astroviral infections commonly cause acute nonbacterial gastroenteritis in children globally. However, these infections often go undiagnosed outside of research settings. There is no treatment available for astrovirus, and Astroviridae strain diversity presents a challenge to potential vaccine development. To address our hypothesis that host genetic risk factors are associated with astrovirus disease susceptibility, we performed a genome-wide association study (GWAS) of astrovirus infection in the first year of life from children enrolled in two Bangladeshi birth cohorts. We identified a novel region on chromosome 1 near the loricrin gene (*LOR*) associated with astrovirus diarrheal infection (rs75437404, meta-analysis p-value=8.82×10^−9^, A allele OR=2.71) and on chromosome 10 near the prolactin releasing hormone receptor gene (*PRLHR*) (rs75935441, meta-analysis p-value=1.33×10^−8^, C allele OR=4.17). The prolactin-releasing peptide has been shown to influence feeding patterns and energy balance in mice. Additionally, several SNPs in the chromosome 1 locus have previously been associated with expression of innate immune system genes *PGLYRP4, S100A9*, and *S100A12*. This study identified two significant host genetic regions that may influence astrovirus diarrhea susceptibility and should be considered in further studies.

## Introduction

Diarrhea is the eighth leading cause of death across all ages and the second leading cause of death among children under 5 years of age (1,2). There were 70.6 deaths per 100 000 children in this age group globally in 2016, compared to 1.3 deaths per 100 000 children within high income countries (3). Classical human astroviruses (HAstV) alone account for 2-9% of all cases of acute nonbacterial gastroenteritis in children across the world, and an analysis of a Bangladeshi birth cohort found that 15-20% of all diarrhea samples in the first year of life were positive for astrovirus (4,5). Astroviral infections are characterized by 2-3 days of watery diarrhea but unlike many other enteric pathogens, astrovirus causes little histological change to intestinal epithelia, including inflammatory responses and cell death (4). The virus can be recovered from the feces of asymptomatic children, suggesting the possibility of prolonged viral shedding, or possibly a mechanism that allows astrovirus to remain in the gastrointestinal tract while causing epithelial barrier dysfunction (6–10). Beyond gastrointestinal infections, there are reported cases of immunocompromised patients with encephalitis and meningitis where astrovirus RNA has been detected in cerebrospinal fluid (10,11). Coupled with other known target organs in animals and its ability to infect across species, the burden of disease associated with astroviral infections may be higher than expected. In patients with gastroenteritis associated astrovirus infections, the only available therapy is fluid replacement to avoid dehydration, as treatment for the virus does not exist.

Astrovirus is most commonly spread from person-to-person, especially via fecal-to-oral transmission and drinking water routes (12,13). There are two types of astrovirus, classic and novel, that are determined by genetic similarity; novel astroviruses are phylogenetically distant from classical HAstVs (11). Both classic and novel astroviruses circulate globally, with classic viruses more prevalent in developing countries (4,10). Astrovirus’s persistence in high-income settings suggests prevention of morbidity requires strategies beyond hygiene improvements (14). Of the three open reading frames (ORFs) in the astrovirus genome, classic astroviruses share 64-84% of capsid amino acid similarities and 93-95% of nucleotide similarities in part of the second ORF (10). Novel astroviruses belong to different clades than classic astroviruses, sharing up to 54% of amino acid identity with classic astroviruses (10). The wide range of genetic diversity within Astroviridae makes potential vaccine development especially difficult.

We lack an understanding of the risk factors associated with astrovirus infections and disease severity. To identify host genetic risk factors associated with astrovirus disease susceptibility that may explain disease mechanism and vaccine targets, we performed genome-wide association studies of children enrolled in two birth cohorts, the Performance of Rotavirus and Oral Polio Vaccines in Developing Countries (PROVIDE) study and the Cryptosporidiosis and Enteropathogens in Bangladesh birth cohort (CBC) in Dhaka, Bangladesh (15,16). We meta-analyzed the results and identified two novel regions on chromosome 1 and 10 significantly associated with astrovirus diarrheal infections in infants.

## Methods

The PROVIDE study protocol was approved by the Research Review Committee (RRC) and Ethics Review Committee (ERC) at the International Centre for Diarrhoeal Disease Bangladesh (icddr,b) and at Institutional Review Boards of the University of Virginia and University of Vermont prior to implementation. The Ethics and Research Review Committee at icddr,b approved the CBC study. For both studies, informed written consent was obtained from the participants or the parents or guardians of all participants.

### PROVIDE Study Design

The “Performance of Rotavirus and Oral Polio Vaccines in Developing Countries” (PROVIDE) study, consisting of children from the Mirpur area of Dhaka, Bangladesh, aimed to evaluate the efficacy of oral and injectable vaccines using a randomized controlled clinical trial 2×2 factorial design (ClinicalTrials.gov registration no. NCT01375647) (15). From 2011-2016, the 700 children in the birth cohort and their mothers were followed for the child’s first two years of life, with biweekly diarrhea surveillance conducted in the homes by field research assistants. Active episodes of diarrhea were referred to the study clinic for evaluation and treatment. For each episode, a diarrhea stool specimen was collected. Height-for-age Z-scores (HAZ) and weight-for-age Z-scores (WAZ) were collected every 3 months.

### CBC Study Design

The Cryptosporidiosis and Enteropathogens in Bangladesh (CBC) study investigated the disease burden of cryptosporidiosis and its effect on children’s growth in urban and rural Bangladesh (ClinicalTrials.gov registration no. NCT02764918) (16). A total of 500 children from Mirpur, Dhaka and 258 children from Mirzapur, a rural subdistrict located near Dhaka, were enrolled at birth. From 2014-2018, twice-weekly in-home visits were conducted by field research assistants to collect data regarding diarrhea and child morbidity, and HAZ and WAZ were collected every 3 months. The study clinic was available to children and caregivers for development of symptoms of any illness. Stool samples were collected monthly and during episodes of diarrhea. Only a subset of children from the Mirpur site had PCR testing of diarrheal samples (n=220) and were therefore included in this study.

### Case and Control Definitions

Stool samples collected in both cohorts were tested for the presence of pathogens via real-time reverse transcription-PCR (RT-PCR) using TaqMan Array Cards (17). Bar graphs displaying the distribution of RT-PCR Ct values for astrovirus in diarrhea samples were plotted using R v3.5.1 (Supp. Fig. 1). Cases attributable to astrovirus were defined as children with diarrheal samples collected within the first year of life that resulted in RT-PCR Ct values for astrovirus >0 and <30. Children were defined as controls if they had at least one diarrhea sample available for testing from the first year of life but all RT-PCR Ct values for astrovirus were ≥30.

### Genotyping Array

In PROVIDE, children were genotyped on the Expanded Multi-Ethnic Genotyping Array (MEGA-EX) from Illumina and children from CBC were genotyped on Illumina’s Infinium Multiethnic Global Array (MEGA). This genetic data was phased with SHAPEIT v2 and imputed with IMPUTE v2.3.2 with 1000 Genomes Project Phase 3 Data as the reference. Standard quality control metrics were used for the genome-wide data. Single nucleotide polymorphism (SNP) filters included genotype missingness <5%, minor allele frequency (MAF) >0.05, and Hardy-Weinberg equilibrium p-value >10^−5^. The PROVIDE cohort had 10 792 283 initial variants and the total genotyping rate was 0.99. A total of 590 340 variants were removed due to missing genotype data, 1 487 431 variants were removed due to minor allele threshold, and 131 variants were removed due to Hardy-Weinberg exact test, leaving 8 777 081 variants. In CBC, there were 10 942 212 initial variants and a total genotyping rate of 0.99. A total of 532 850 variants were lost to missing genotype data, 1 528 417 were removed due to minor allele threshold, and 8 were removed due to Hardy-Weinberg exact test, resulting in 8 880 118 variants that passed the quality control filters. There were 18 individuals in CBC who were identified as outliers in the principal components analysis based on their Principal Component (PCA) score. Those with PCA scores falling 3 standard deviations above and below the median PCA value were removed (Supp. Fig. 2a, 2b). The genomic inflation factor, or λ, showed no inflation, λ=1.003 and 1.033 for PROVIDE and CBC, respectively (Supp. Fig. 3).

### Association Analysis

Genome-wide association analyses using homo sapiens (human) genome assembly GRCh37 (hg19) from the Genome Reference Consortium were performed separately for each study using logistic regression with an additive model within SNPTEST v2. Manhattan plots were constructed for each cohort using the ggplot2 package in R v.3.6.1 and highlights of regions of interest were created using LocusZoom tools at locuszoom.org with hg19/1000 Genomes SAS as the reference genome build.

Data from PROVIDE and CBC were combined for a fixed-effect meta-analysis using METAL (18). Input from both cohorts was filtered on MAF > 5% and IMPUTE2 (INFO) > 0.7, retaining SNPs that had been imputed with high certainty in the dataset. Functional Mapping and Annotation of Genome-Wide Association Studies (FUMA) was used to annotate genomic regions of interest and identify variants in linkage disequilibrium (19). This annotation included expression quantitative trait loci (eQTL) from several databases, including the Genotype-Tissue Expression project (GTEx) and the eQTLGen Consortium (20). Conditional analyses were performed at each associated locus using SNPTEST v2 in each cohort and then combined for an overall fixed-effect meta-analysis in METAL.

## Results

Within PROVIDE, 119 children were identified as having at least one astrovirus associated diarrheal case in the first year of life and 314 children did not have an associated astroviral infection. Children with astrovirus associated diarrhea had an earlier mean age (103.9 days) of first diarrheal episode as compared to children without astrovirus infection (136.7 days). Similarly, there were 58 astrovirus associated diarrheal cases and 96 controls in CBC, with a mean age of first diarrheal episode of 117.7 days for cases and 125.2 days for controls. In both cohorts, children not identified as cases or controls either left the study early, did not have diarrhea, or had one or more samples with missing data for astrovirus. The distribution of HAZ and WAZ did not differ between cases and controls in either cohort at birth or at 12 months of age (Table 1). Cases and controls both had a mean mild diarrheal severity score in PROVIDE and a mean moderate diarrheal severity score in CBC (Table 1).

**Table 1.**
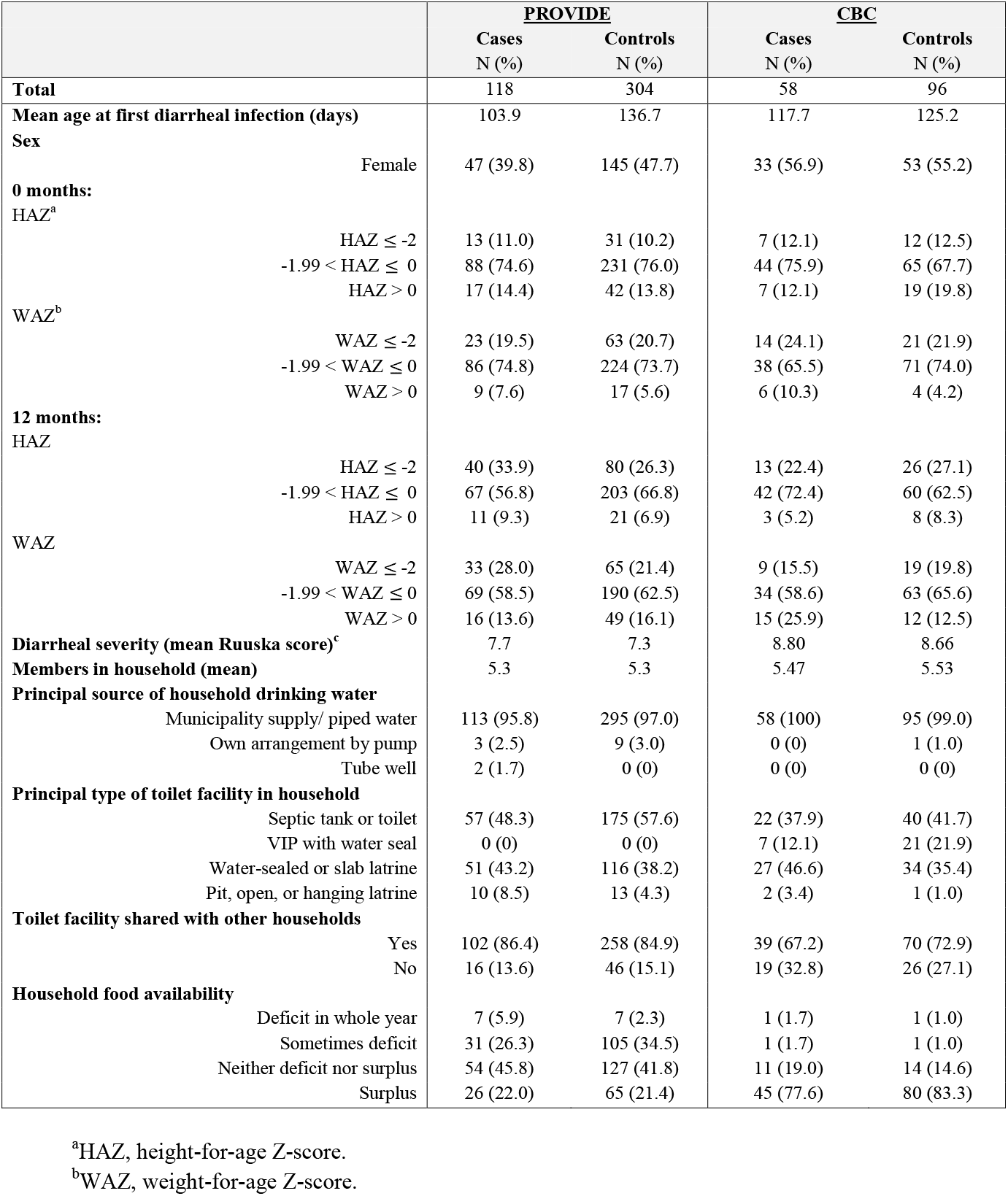
Characteristics of children in PROVIDE and CBC cohorts.

|  | <u>PROVIDE</u> |  | <u>CBC</u> |  |
| --- | --- | --- | --- | --- |
|  | Cases<br>N (%) | Controls<br>N (%) | Cases<br>N (%) | Controls<br>N (%) |
| **Total** | 118 | 304 | 58 | 96 |
| **Mean age at first diarrheal infection (days)** | 103.9 | 136.7 | 117.7 | 125.2 |
| **Sex** |  |  |  |  |
| Female | 47 (39.8) | 145 (47.7) | 33 (56.9) | 53 (55.2) |
| **0 months:** |  |  |  |  |
| HAZ<sup>a</sup> |  |  |  |  |
| HAZ ≤ -2 | 13 (11.0) | 31 (10.2) | 7 (12.1) | 12 (12.5) |
| -1.99 < HAZ ≤ 0 | 88 (74.6) | 231 (76.0) | 44 (75.9) | 65 (67.7) |
| HAZ > 0 | 17 (14.4) | 42 (13.8) | 7 (12.1) | 19 (19.8) |
| WAZ<sup>b</sup> |  |  |  |  |
| WAZ ≤ -2 | 23 (19.5) | 63 (20.7) | 14 (24.1) | 21 (21.9) |
| -1.99 < WAZ ≤ 0 | 86 (74.8) | 224 (73.7) | 38 (65.5) | 71 (74.0) |
| WAZ > 0 | 9 (7.6) | 17 (5.6) | 6 (10.3) | 4 (4.2) |
| **12 months:** |  |  |  |  |
| HAZ |  |  |  |  |
| HAZ ≤ -2 | 40 (33.9) | 80 (26.3) | 13 (22.4) | 26 (27.1) |
| -1.99 < HAZ ≤ 0 | 67 (56.8) | 203 (66.8) | 42 (72.4) | 60 (62.5) |
| HAZ > 0 | 11 (9.3) | 21 (6.9) | 3 (5.2) | 8 (8.3) |
| WAZ |  |  |  |  |
| WAZ ≤ -2 | 33 (28.0) | 65 (21.4) | 9 (15.5) | 19 (19.8) |
| -1.99 < WAZ ≤ 0 | 69 (58.5) | 190 (62.5) | 34 (58.6) | 63 (65.6) |
| WAZ > 0 | 16 (13.6) | 49 (16.1) | 15 (25.9) | 12 (12.5) |
| **Diarrheal severity (mean Ruuska score)<sup>c</sup>** | 7.7 | 7.3 | 8.80 | 8.66 |
| **Members in household (mean)** | 5.3 | 5.3 | 5.47 | 5.53 |
| **Principal source of household drinking water** |  |  |  |  |
| Municipality supply/ piped water | 113 (95.8) | 295 (97.0) | 58 (100) | 95 (99.0) |
| Own arrangement by pump | 3 (2.5) | 9 (3.0) | 0 (0) | 1 (1.0) |
| Tube well | 2 (1.7) | 0 (0) | 0 (0) | 0 (0) |
| **Principal type of toilet facility in household** |  |  |  |  |
| Septic tank or toilet | 57 (48.3) | 175 (57.6) | 22 (37.9) | 40 (41.7) |
| VIP with water seal | 0 (0) | 0 (0) | 7 (12.1) | 21 (21.9) |
| Water-sealed or slab latrine | 51 (43.2) | 116 (38.2) | 27 (46.6) | 34 (35.4) |
| Pit, open, or hanging latrine | 10 (8.5) | 13 (4.3) | 2 (3.4) | 1 (1.0) |
| **Toilet facility shared with other households** |  |  |  |  |
| Yes | 102 (86.4) | 258 (84.9) | 39 (67.2) | 70 (72.9) |
| No | 16 (13.6) | 46 (15.1) | 19 (32.8) | 26 (27.1) |
| **Household food availability** |  |  |  |  |
| Deficit in whole year | 7 (5.9) | 7 (2.3) | 1 (1.7) | 1 (1.0) |
| Sometimes deficit | 31 (26.3) | 105 (34.5) | 1 (1.7) | 1 (1.0) |
| Neither deficit nor surplus | 54 (45.8) | 127 (41.8) | 11 (19.0) | 14 (14.6) |
| Surplus | 26 (22.0) | 65 (21.4) | 45 (77.6) | 80 (83.3) |

<sup>a</sup>HAZ, height-for-age Z-score.
<sup>b</sup>WAZ, weight-for-age Z-score.

### Genetic Associations

Figure 1 shows the association results for each cohort and the meta-analysis. The top association (rs75437404, meta-analysis p-value=8.82×10^−9^, MAF=16.7%) was identified in a non-coding region on chromosome 1 near the loricrin gene (*LOR*) (Table 2). The *LOR* gene consists of 2.4 Kb, and the peak association region around this SNP spans 88 Kb (Fig. 2a) (21). Children with at least 1 copy of the A allele at SNP rs75437404 were 2.7 times as likely to have an astrovirus associated diarrheal infection within the first year of life compared to children with the T allele. This top associated SNP, rs75437404, has a MAF of 17.2% in PROVIDE and 15.2% in CBC, and is in linkage disequilibrium (LD, r^2^>0.8) with 13 variants spanning an intergenic region 15.3 Kb upstream of *LOR*. A conditional analysis incorporating rs75437404 in the model attenuated the association, suggesting the region is in strong linkage disequilibrium and the alleles are correlated (Fig. 2c).

**Figure 1.**
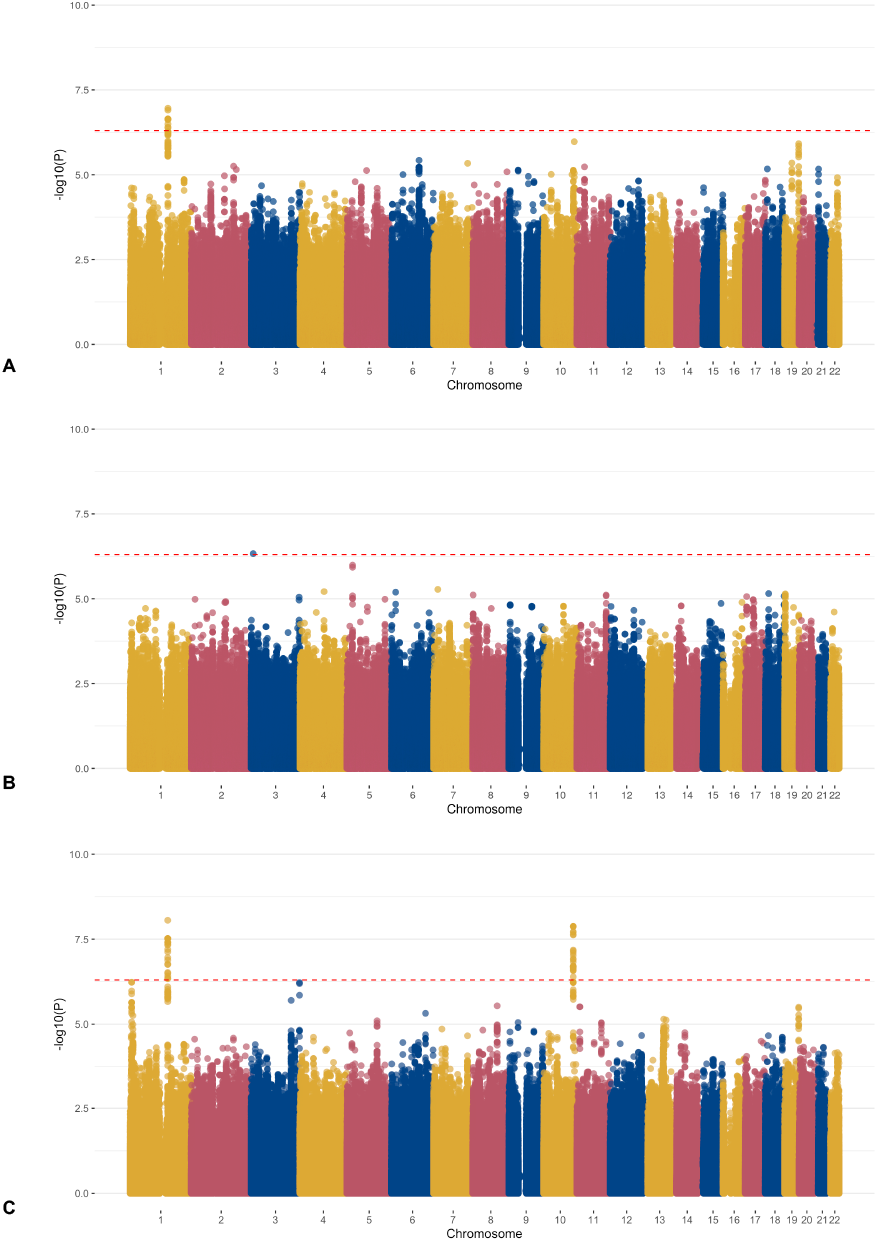
SNP associations with astrovirus associated diarrhea in the first year of life. Manhattan plots showing -log_10_ p-values for SNP associations by cohort. Dashed line is threshold for genome-wide significance (5×10^−7^). **A)** PROVIDE. **B)** CBC. **C)** Meta-analysis.

**Figure 2.**
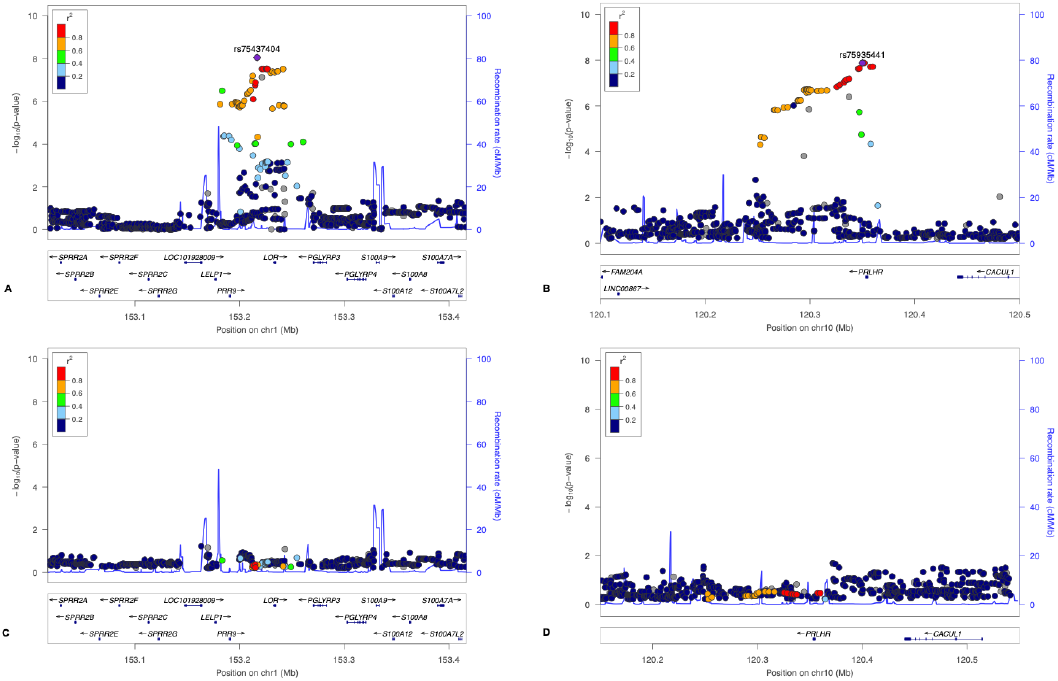
Regional association plots for two meta-analysis loci **a)** Chromosome 1: *LOR* region. **b)** Chromosome 10: *PRLHR* region. **c)** Chromosome 1: *LOR* region conditioning on rs75437404. **d)** Chromosome 10: *PRLHR* region conditioning on rs75935441.

**Table 2.** Genome-wide significant associated SNPs from stratified analyses and meta-analysis.

| SNP/Variant | Chr<sup>a</sup> | Position | <u>PROVIDE (N=433)</u> |  |  | <u>CBC (N=154)</u> |  |  | <u>Meta-Analysis</u> |  |  |
| --- | --- | --- | --- | --- | --- | --- | --- | --- | --- | --- | --- |
|  |  |  | OR<sup>b</sup> | MAF<sup>c</sup> | p-value | OR | MAF | p-value | OR | 95% CI<sup>d</sup> | p-value |
| rs75437404 | 1 | 153216817 | 2.60 | 0.17 | $1.09 \times 10^{-7}$ | 2.12 | 0.15 | $1.81 \times 10^{-2}$ | 2.71 | 1.93-3.81 | $8.82 \times 10^{-9}$ |
| rs12131158 | 1 | 153221315 | 2.53 | 0.19 | $2.28 \times 10^{-7}$ | 1.94 | 0.17 | $2.99 \times 10^{-2}$ | 2.47 | 1.79-3.40 | $3.02 \times 10^{-8}$ |
| rs7518709 | 1 | 153225883 | 2.53 | 0.19 | $2.28 \times 10^{-7}$ | 1.94 | 0.17 | $2.99 \times 10^{-2}$ | 2.47 | 1.79-3.40 | $3.02 \times 10^{-8}$ |
| rs7542150 | 1 | 153225921 | 2.53 | 0.19 | $2.27 \times 10^{-7}$ | 1.94 | 0.17 | $2.99 \times 10^{-2}$ | 2.47 | 1.79-3.40 | $3.01 \times 10^{-8}$ |
| rs6666892 | 1 | 153226579 | 2.53 | 0.19 | $2.28 \times 10^{-7}$ | 1.94 | 0.17 | $2.99 \times 10^{-2}$ | 2.47 | 1.79-3.40 | $3.02 \times 10^{-8}$ |
| rs6661601 | 1 | 153233510 | 2.39 | 0.20 | $6.22 \times 10^{-7}$ | 2.02 | 0.17 | $1.89 \times 10^{-2}$ | 2.44 | 1.77-3.36 | $4.46 \times 10^{-8}$ |
| rs12408942 | 1 | 153241677 | 2.44 | 0.20 | $3.95 \times 10^{-7}$ | 2.01 | 0.17 | $1.96 \times 10^{-2}$ | 2.47 | 1.79-3.41 | $3.06 \times 10^{-8}$ |
| rs114810342 | 10 | 120346096 | 2.91 | 0.07 | $1.60 \times 10^{-5}$ | 4.82 | 0.07 | $2.80 \times 10^{-4}$ | 4.05 | 2.48-6.62 | $2.38 \times 10^{-8}$ |
| rs55907016 | 10 | 120347235 | 2.91 | 0.07 | $1.54 \times 10^{-5}$ | 4.81 | 0.07 | $3.96 \times 10^{-4}$ | 4.06 | 2.48-6.63 | $2.30 \times 10^{-8}$ |
| rs75935441 | 10 | 120350104 | 2.99 | 0.07 | $7.71 \times 10^{-6}$ | 4.58 | 0.07 | $3.96 \times 10^{-4}$ | 4.17 | 2.55-6.83 | $1.33 \times 10^{-8}$ |
| rs116216801 | 10 | 120350154 | 2.99 | 0.07 | $7.71 \times 10^{-6}$ | 4.58 | 0.07 | $3.96 \times 10^{-4}$ | 4.17 | 2.55-6.83 | $1.33 \times 10^{-8}$ |
| rs115109981 | 10 | 120350780 | 2.99 | 0.07 | $7.70 \times 10^{-6}$ | 4.57 | 0.07 | $3.71 \times 10^{-4}$ | 4.17 | 2.55-6.83 | $1.34 \times 10^{-8}$ |
| rs117647802 | 10 | 120351875 | 2.99 | 0.07 | $7.69 \times 10^{-6}$ | 4.57 | 0.07 | $3.72 \times 10^{-4}$ | 4.17 | 2.55-6.82 | $1.34 \times 10^{-8}$ |
| rs116989176 | 10 | 120357518 | 3.00 | 0.08 | $7.43 \times 10^{-6}$ | 4.22 | 0.07 | $6.19 \times 10^{-3}$ | 4.08 | 2.50-6.66 | $1.95 \times 10^{-8}$ |
| rs1711891 | 10 | 120359829 | 3.00 | 0.08 | $7.42 \times 10^{-6}$ | 4.23 | 0.07 | $6.18 \times 10^{-3}$ | 4.07 | 2.50-6.65 | $1.93 \times 10^{-8}$ |

<sup>a</sup>Chr, chromosome.<sup>b</sup>OR, odds ratio.<sup>c</sup>MAF, minor allele frequency.<sup>d</sup>CI, confidence interval.

We also identified another region, including the prolactin releasing hormone receptor gene (*PRLHR*). The peak association region around *PRLHR* spans 51 Kb, including the 9.9 Kb gene (Fig. 2b) on chromosome 10 (Fig. 1a, 1c) (21). SNP rs75935441 is in the 3’ untranslated region of the *PRLHR* gene (meta-analysis p-value=1.33×10^−8^, MAF=7.5%) and is part of an enhancer sequence (22). Children with at least 1 copy of the C allele at rs75935441 were 4 times as likely to have an astrovirus diarrheal infection compared to children with the T allele (OR=4.17, 95% CI=2.55-6.83). The conditional analysis including rs75935441 in the model attenuated the signal (Fig. 2d).

Using FUMA, we identified chromatin interactions with the promoter regions of 55 genes on chromosome 1, including *LOR, PRR9, PGLYRP4, S100A9* and *S100A12* (Fig. 3a, Supp. Table 1) spanning base pairs (bp) 153 181 362 – 153 242 630. Additionally, 11 genes had one or more expression quantitative trait loci (eQTLs) in this region (Supp. Table 2). Several variants have been previously associated with expression of multiple genes. For example, rs12125683 (C allele OR=2.23, meta-analysis p-value=7.92×10^−7^) is associated with expression of *PGLYRP4, S100A12*, and *S100A9*. The C allele of the SNP is associated with higher expression of *PGLYRP4* in esophagus mucosa (p-value=2.69×10^−5^), as well as higher expression of *S100A12* (p-value=1.94×10^−23^) and *S100A9* (p-value=1.27×10^−6^) in whole blood (Supp. Table 2).

**Figure 3.**
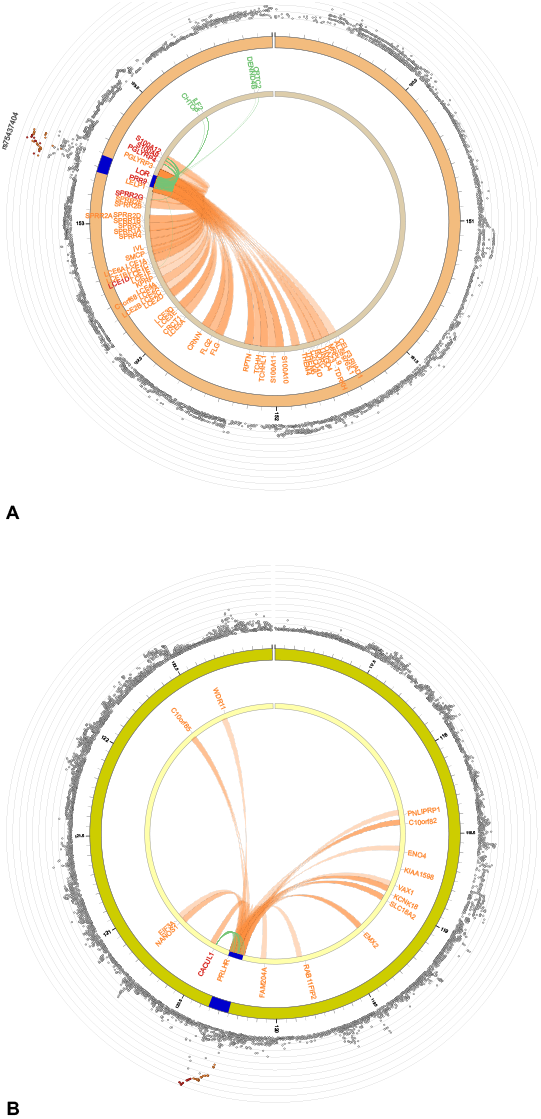
Circos plots of chromatin interactions. The outermost ring shows the results from the GWAS meta-analysis for **(A)** chromosome 1 and **(B)** chromosome 10. The next ring shows base pair coordinates along the chromosome (in Mb), with the region of interest colored blue. The innermost ring is annotated with gene symbols for all genes that had either a chromatin interaction (orange), an eQTL (green), or both (red).

Similarly, the chromosome 10 region (bp 120 252 606 – 120 359 829) had chromatin interactions with the promoter regions of 16 genes on chromosome 10, including *PRLHR, CACUL1, KCNK18, ENO4*, and *EIF3A* (Fig. 3b, Supp. Table 1). Only 1 gene also had identified eQTLs: *CACUL1*. The top associated SNP in this locus, rs75935441 (C allele OR=4.17, meta-analysis p-value=1.33×10^−8^), is an eQTL for *CACUL1* and the C allele is associated with lower expression in both whole blood (p-value=1.20×10^−12^) and skeletal muscle (p-value=4.36×10^−6^) (Supp. Table 2).

## Discussion

We identified two significant regions across the genome that were associated with astrovirus diarrheal infection in the first year of life. The first region is upstream of the *LOR* gene and is associated with expression of immune system genes *PGLYRP4, S100A12*, and *S100A9*. The second region encompasses the *PLRHR* gene and is associated with expression of the gene *CACUL1*.

*LOR* encodes loricrin, which contributes to the protective barrier function of the epidermis, and is expressed almost exclusively in mammalian stratified epithelia (23). It is also found in macrophages in various tissues (24), but the relationship to astrovirus is not evident. However, within the chromosome 1 locus, the presence of chromatin interactions coupled with identification of eQTLs indicates that several of these variants are likely regulatory in nature. Both *S100A12* and *S100A9* are in the ‘MyD88 dependent cascade initiated on endosome’ SuperPath (21). Astrovirus enters cells via endocytosis and when toll-like receptors bind viral nucleic acid inside the cell, they initiate signaling cascades which include MyD88 and type 1 IFN (25,26). The S100A9 protein regulates toll-like receptor 3 activation (27,28) and the S100A12 protein is an endogenous activator of toll-like receptor 4 (29). Both proteins are calgranulins and in addition to TLR activation are also involved in protecting the body against damage from inflammation (30,31). The same allele associated with higher expression of these genes in whole blood is associated with increased odds of astrovirus positive diarrhea in the first year of life. The C allele of rs12125683 is also associated with higher expression of *PGLYRP4* in esophagus mucosa. *PGLYRP4* is a peptidoglycan recognition protein, involved in the innate immune response to bacteria (21). Higher expression of this gene could contribute to astrovirus susceptibility via changes to the gastrointestinal microbiome (32).

The *PRLHR* gene is also in a region significantly associated with astrovirus diarrheal infections. This gene encodes the receptor for prolactin-releasing peptide, which has been identified as a target for obesity treatment (33). In mouse and rat models, prolactin-releasing peptide influences feeding patterns and energy balance, and it has shown diet-suppressing effects (33,34). However, we further stratified by WAZ and HAZ to account for any underlying malnutrition at birth or at 12 months of age that may predispose an individual to an infection, and there were no differences between cases and controls (Table 1). Thus, the association between *PRLHR* and astrovirus diarrhea infections is not likely mediated by malnutrition. Further investigation to identify the potential association between *PRLHR* and the astrovirus replication cycle or its pathogenic mechanism is warranted.

In addition to the effects on *PRLHR*, we found SNPs associated with increased risk of astrovirus positive diarrhea that were also associated with lower expression of the gene *CACUL1* in whole blood. This gene promotes cell proliferation and higher expression has been linked to increased invasion of gastric cancer cells (35). While linked to *Helicobacter pylori* activity (35), it is unclear how this gene may be involved in astrovirus pathogenesis. Additional chromatin interactions at this locus suggest the potential for regulation of the genes *ENO4, KCNK18*, and *EIF3A*, but there are not yet data showing differences in gene expression based on genotypes found in the region of interest.

In conclusion, we identified two genetic regions associated with astrovirus infection susceptibility. Both regions warrant additional exploration for their potential association with immune and gastric cells and may elucidate astrovirus infection pathways.

## Data Availability

Data are publicly available from the NIH, via dbGAP, phs001478.v1.p1 and phs001665.v1.p1, or by request from us.

## Acknowledgments

We would like to thank the families that participated in these studies, as well as the field research assistants and laboratory staff. icddr,b is grateful to the governments of Bangladesh, Canada, Sweden, and the UK for their unrestricted core support.

## Author Contributions

RH, WAP, and PD conceived and designed the study. RH, AJM, BDK, and WAP acquired data. RH, UN, GW, AJM, BDK, WAP, and PD curated data. LC, RMM, DD, and PD analyzed and interpreted data, with assistance in interpretation from GW, PK, and WAP. All authors were responsible for writing the manuscript and approved the final version for publication.

## References

1. The top 10 causes of death [Internet]. [cited 2023 Mar 12]. Available from: https://www.who.int/news-room/fact-sheets/detail/the-top-10-causes-of-death

2. Diarrhoea [Internet]. [cited 2023 Mar 12]. Available from: https://www.who.int/health-topics/diarrhoea

3. Limaheluw J, Medema G, Hofstra N. An exploration of the disease burden due to Cryptosporidium in consumed surface water for sub-Saharan Africa. Int J Hyg Environ Health. 2019 Jun 1;222(5):856–63.

4. Bosch A, Pintó RM, Guix S. Human Astroviruses. Clin Microbiol Rev. 2014 Oct;27(4):1048–74.

5. Schnee AE, Haque R, Taniuchi M, Uddin MJ, Alam MM, Liu J, et al. Identification of Etiology-Specific Diarrhea Associated With Linear Growth Faltering in Bangladeshi Infants. Am J Epidemiol. 2018 Oct 1;187(10):2210–8.

6. Resque HR, Munford V, Castilho JG, Schmich H, Caruzo TAR, Rácz ML. Molecular characterization of astrovirus in stool samples from children in São Paulo, Brazil. Mem Inst Oswaldo Cruz. 2007 Dec;102:969–74.

7. Kapusinszky B, Minor P, Delwart E. Nearly Constant Shedding of Diverse Enteric Viruses by Two Healthy Infants. J Clin Microbiol. 2012 Nov;50(11):3427–34.

8. Chhabra P, Payne DC, Szilagyi PG, Edwards KM, Staat MA, Shirley SH, et al. Etiology of Viral Gastroenteritis in Children <5 Years of Age in the United States, 2008–2009. J Infect Dis. 2013 Sep 1;208(5):790–800.

9. Meyer CT, Bauer IK, Antonio M, Adeyemi M, Saha D, Oundo JO, et al. Prevalence of classic, MLB-clade and VA-clade Astroviruses in Kenya and The Gambia. Virol J. 2015 May 15;12:78.

10. Vu DL, Bosch A, Pintó RM, Guix S. Epidemiology of Classic and Novel Human Astrovirus: Gastroenteritis and Beyond. Viruses. 2017 Feb 18;9(2):33.

11. Vu DL, Cordey S, Brito F, Kaiser L. Novel human astroviruses: Novel human diseases? J Clin Virol. 2016 Sep 1;82:56–63.

12. Kurtz JB, Lee TW, Craig JW, Reed SE. Astrovirus infection in volunteers. J Med Virol. 1979;3(3):221–30.

13. Abad FX, Pintó RM, Villena C, Gajardo R, Bosch A. Astrovirus survival in drinking water. Appl Environ Microbiol. 1997 Aug;63(8):3119–22.

14. Olortegui MP, Rouhani S, Yori PP, Salas MS, Trigoso DR, Mondal D, et al. Astrovirus Infection and Diarrhea in 8 Countries. Pediatrics. 2018 Jan;141(1):e20171326.

15. Kirkpatrick BD, Colgate ER, Mychaleckyj JC, Haque R, Dickson DM, Carmolli MP, et al. The “Performance of Rotavirus and Oral Polio Vaccines in Developing Countries” (PROVIDE) study: description of methods of an interventional study designed to explore complex biologic problems. Am J Trop Med Hyg. 2015 Apr 1;92(4):744–51.

16. Steiner KL, Ahmed S, Gilchrist CA, Burkey C, Cook H, Ma JZ, et al. Species of Cryptosporidia causing subclinical infection associated with growth faltering in rural and urban Bangladesh: a birth cohort study. Clin Infect Dis. 2018 Oct 15;67(9):1347–55.

17. Liu J, Gratz J, Amour C, Kibiki G, Becker S, Janaki L, et al. A laboratory-developed TaqMan Array Card for simultaneous detection of 19 enteropathogens. J Clin Microbiol. 2013 Feb 1;51(2):472–80.

18. Willer CJ, Li Y, Abecasis GR. METAL: fast and efficient meta-analysis of genomewide association scans. Bioinformatics. 2010 Sep 1;26(17):2190–1.

19. Watanabe K, Taskesen E, van Bochoven A, Posthuma D. Functional mapping and annotation of genetic associations with FUMA. Nat Commun. 2017 Nov 28;8(1):1826.

20. Võsa U, Claringbould A, Westra HJ, Bonder MJ, Deelen P, Zeng B, et al. Large-scale cis- and trans-eQTL analyses identify thousands of genetic loci and polygenic scores that regulate blood gene expression. Nat Genet. 2021 Sep;53(9):1300–10.

21. Stelzer G, Rosen N, Plaschkes I, Zimmerman S, Twik M, Fishilevich S, et al. The GeneCards suite: from gene data mining to disease genome sequence analyses. Curr Protoc Bioinforma. 2016;54(1):1.30.1-1.30.33.

22. Zerbino DR, Wilder SP, Johnson N, Juettemann T, Flicek PR. The Ensembl Regulatory Build. Genome Biol. 2015 Mar 24;16(1):56.

23. Hohl D, Lichti U, Breitkreutz D, Steinert PM, Roop DR. Transcription of the Human Loricrin Gene In Vitro Is Induced by Calcium and Cell Density and Suppressed by Retinoic Acid. J Invest Dermatol. 1991 Apr 1;96(4):414–8.

24. Uhlén M, Fagerberg L, Hallström BM, Lindskog C, Oksvold P, Mardinoglu A, et al. Proteomics. Tissue-based map of the human proteome. Science. 2015 Jan 23;347(6220):1260419.

25. Lester SN, Li K. Toll-Like Receptors in Antiviral Innate Immunity. J Mol Biol. 2014 Mar 20;426(6):1246–64.

26. Marvin SA. The Immune Response to Astrovirus Infection. Viruses. 2016 Dec 30;9(1):1.

27. Tsai SY, Segovia JA, Chang TH, Morris IR, Berton MT, Tessier PA, et al. DAMP Molecule S100A9 Acts as a Molecular Pattern to Enhance Inflammation during Influenza A Virus Infection: Role of DDX21-TRIF-TLR4-MyD88 Pathway. PLOS Pathog. 2014 Jan 2;10(1):e1003848.

28. Tsai SY, Segovia JA, Chang TH, Shil NK, Pokharel SM, Kannan TR, et al. Regulation of toll-like receptor 3 activation by S100A9. J Immunol Baltim Md 1950. 2015 Nov 1;195(9):4426–37.

29. Kessel C, Fühner S, Brockmeyer S, Wittkowski H, Föll D. Hexameric S100A12 is required for pro-inflammatory TLR4-signalling. Pediatr Rheumatol Online J. 2015 Sep 28;13(Suppl 1):O30.

30. Hsu K, Champaiboon C, Guenther BD, Sorenson BS, Khammanivong A, Ross KF, et al. ANTI-INFECTIVE PROTECTIVE PROPERTIES OF S100 CALGRANULINS. Anti-Inflamm Anti-Allergy Agents Med Chem. 2009 Dec 4;8(4):290–305.

31. Jukic A, Bakiri L, Wagner EF, Tilg H, Adolph TE. Calprotectin: from biomarker to biological function. Gut. 2021 Oct;70(10):1978–88.

32. Dabrowski AN, Shrivastav A, Conrad C, Komma K, Weigel M, Dietert K, et al. Peptidoglycan Recognition Protein 4 Limits Bacterial Clearance and Inflammation in Lungs by Control of the Gut Microbiota. Front Immunol. 2019 Sep;10:2106.

33. Kuneš J, Pražienková V, Popelová A, Mikulášková B, Zemenová J, Maletínská L. Prolactin-releasing peptide: a new tool for obesity treatment. J Endocrinol. 2016 Aug;230(2):R51–58.

34. Mikulášková B, Zemenová J, Pirník Z, Pražienková V, Bednárová L, Železná B, et al. Effect of palmitoylated prolactin-releasing peptide on food intake and neural activation after different routes of peripheral administration in rats. Peptides. 2016 Jan;75:109–17.

35. Kong Y, Ma L qing, Bai P song, Da R, Sun H, Qi X gai, et al. Helicobacter pylori promotes invasion and metastasis of gastric cancer cells through activation of AP-1 and up-regulation of CACUL1. Int J Biochem Cell Biol. 2013 Nov;45(11):2666–78.

