## Supplemental Figure 1 for "Genetic Susceptibility to Astrovirus Diarrhea in Bangladeshi Infants"

**a.**

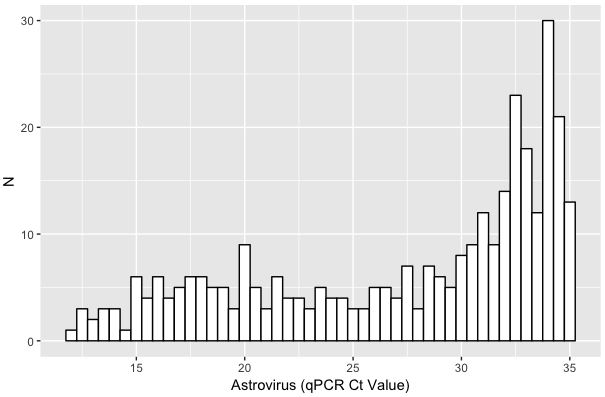

**b.**

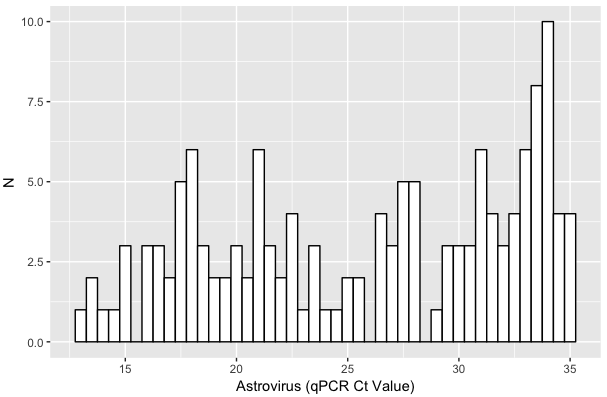

### **Supplementary Figure 1.** Distribution of astroviral load among diarrheal events by cohort. Bar graphs showing frequency of each Ct value, produced in R v3.5.1. **a)** PROVIDE. **b)** CBC.
