## Supplemental Figure 2 for "Genetic Susceptibility to Astrovirus Diarrhea in Bangladeshi Infants"

**a)**

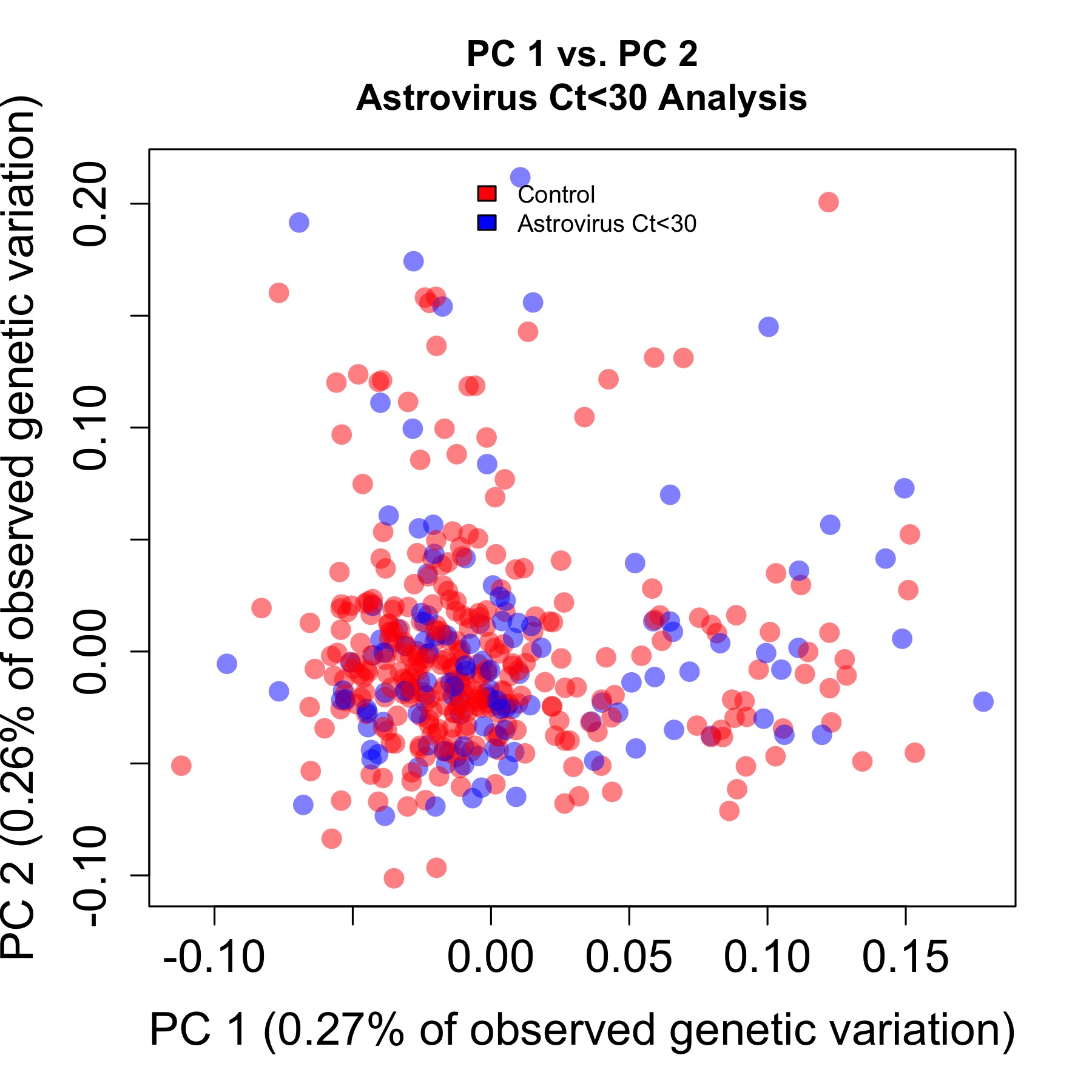

**b)**

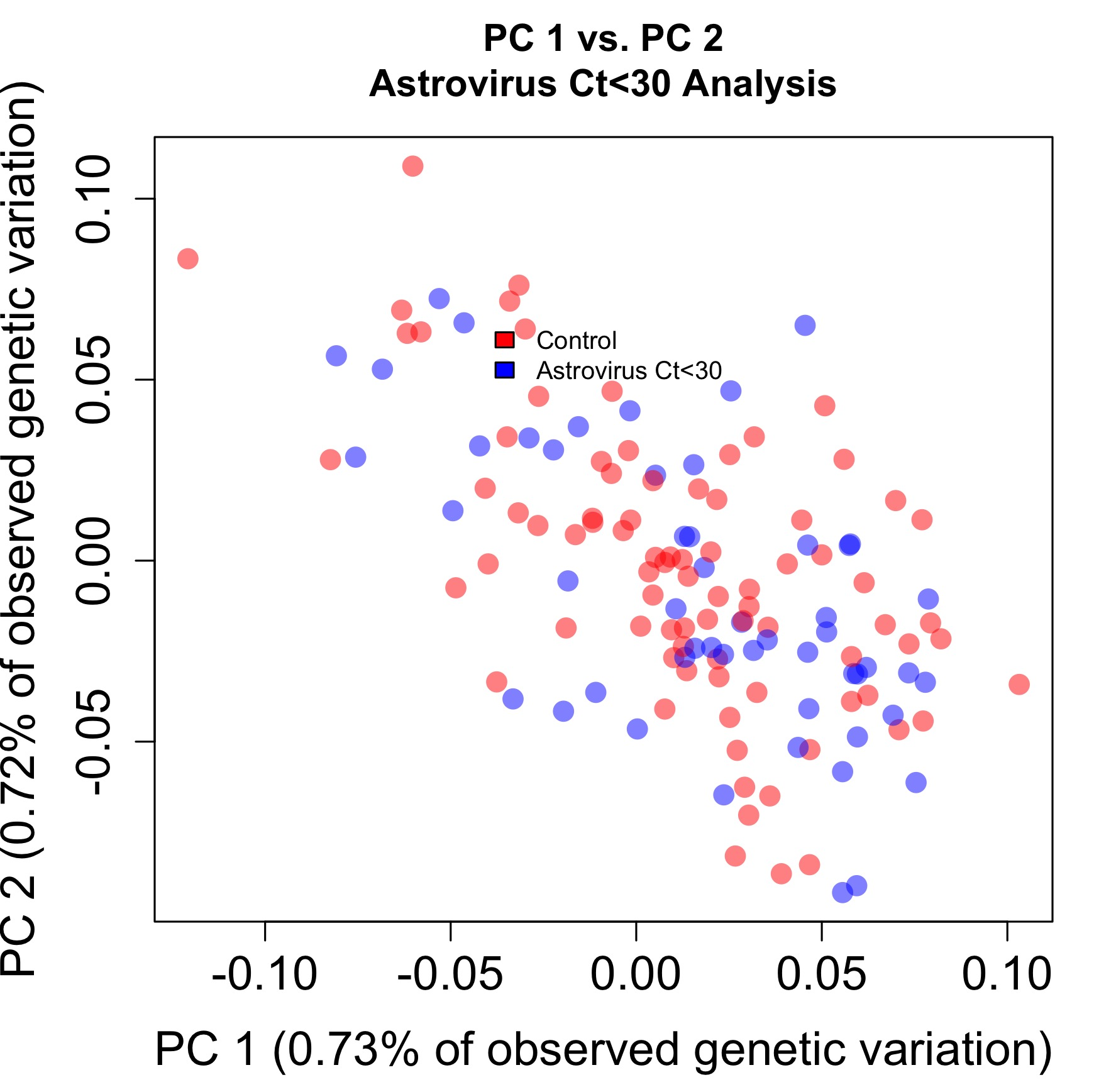

**Supplementary Figure 2.** Principal components analyses. **a)** PROVIDE: PC 1 vs. PC 2. **b)** CBC with outliers removed: PC 1 vs. PC 2.
