## Supplementary figures and images for "Genetic Susceptibility to Astrovirus Diarrhea in Bangladeshi Infants"

### Supplemental Figure 3

**a)**

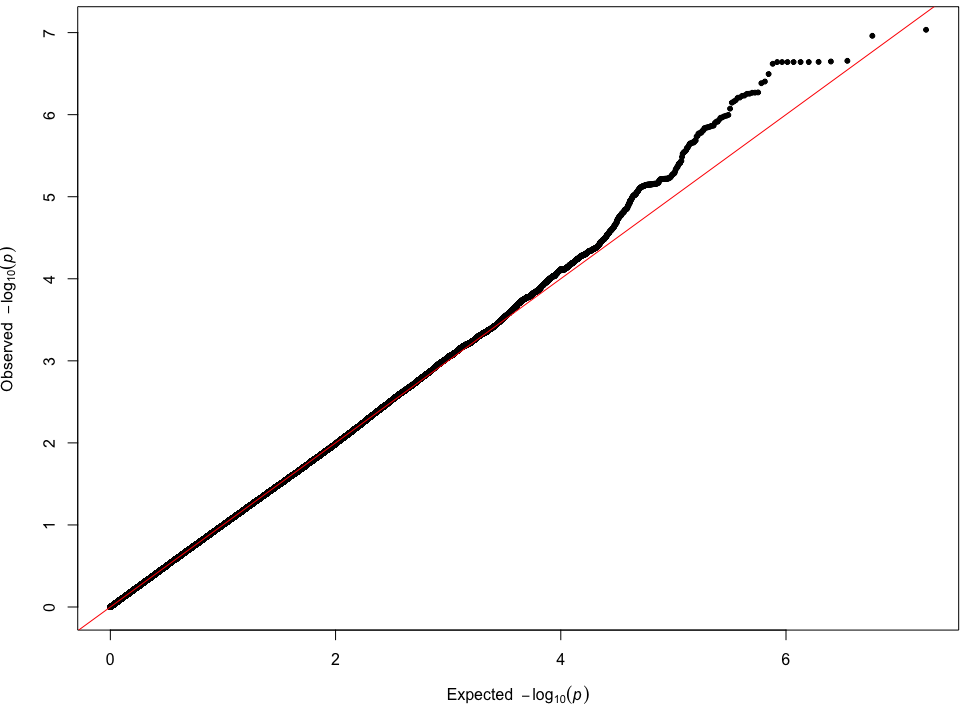

**b)**

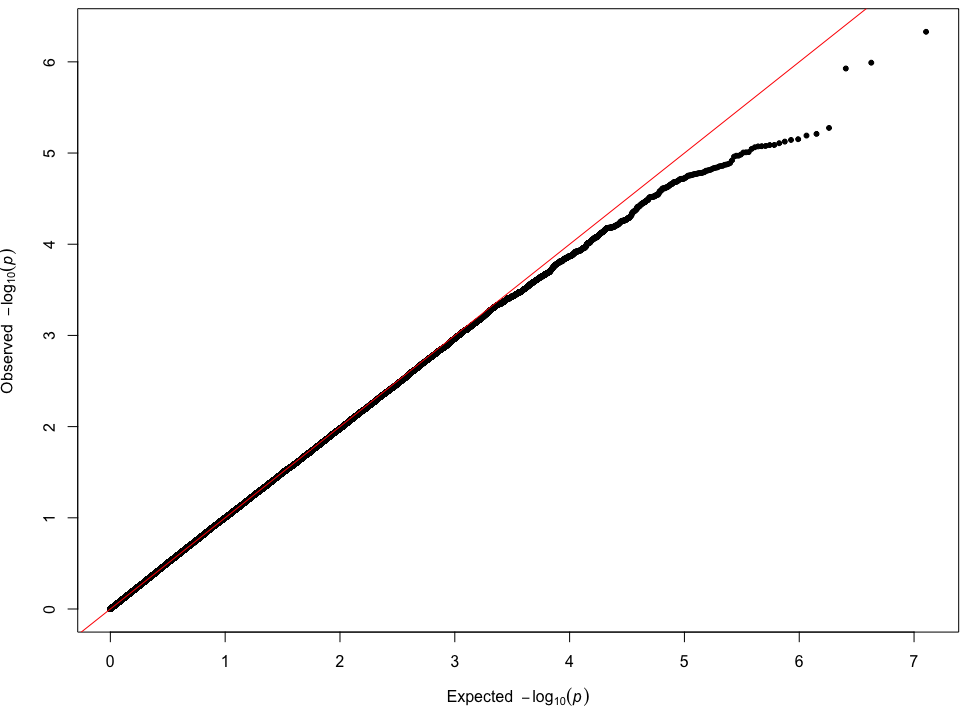

**Supplementary Figure 3.** Quantile-quantile plots of each cohort. **a)** PROVIDE, λ=1.003. **b)** CBC, λ=1.033.
